# Sleep and circadian biomarkers of postoperative delirium: protocol for a prospective, observational cohort study

**DOI:** 10.1101/2023.09.21.23295920

**Authors:** Elizabeth Sugg, Elizabeth Gleeson, Sarah N. Baker, Peng Li, Chenlu Gao, Ariel Mueller, Hao Deng, Shiqian Shen, Esteban Franco Garcia, Richa Saxena, Erik Musiek, Oluwaseun Akeju, Zhongcong Xie, Kun Hu, Lei Gao

**Author notes:** **Correspondence to:** Dr. Lei Gao. Co-first authors (Elizabeth Sugg, Elizabeth Gleeson).

## Abstract

Surgical patients over 70 experience postoperative delirium (POD) complications in up to 50% of procedures. Sleep/circadian disruption has emerged as a potential risk factor for POD in epidemiological studies. However, the relationship remains unclear in a clinical setting. This protocol presents a single-site, prospective, noninvasive observational study designed to examine the relationship between sleep/circadian regulation and POD and how this association is influenced by AD pathology and genetic risk for AD. This novel approach for understanding modifiable risk factors of POD and cognitive decline after surgery could assist with establishing treatments and preventative measures for POD in the future.

## INTRODUCTION

Postoperative delirium (POD) is the most common surgical complication in older adults, occurring in 5% to 50% of cases.[1] POD is a memory and thinking deficits syndrome associated with cognitive decline and Alzheimer’s disease (AD) and related dementias (ADRD). Millions of older Americans require major surgery each year and are exposed to the risks of POD and its consequences. Furthermore, there are currently no treatments for POD, but it perpetuates a cascade of poor health outcomes costing $50 billion annually.[2] Older age, cognitive impairment, and pre-existing AD/ADRD are major risk factors, but POD also predisposes to AD/ADRD, suggesting overlapping pathophysiology. Yet, minimal attention or cognitive follow-up exists for those who suffer from POD.[3] Given that nearly 40% of POD is preventable with attention to multifactorial preoperative care,[4] the search for modifiable risk factors and novel mechanisms in POD is of great public health importance.

At the same time, there is a silent epidemic of treatable chronic sleep problems and circadian (our “body clocks”) disruption, affecting over 70 million older Americans (e.g., insufficient, irregular timing, unscheduled naps, insomnia, or sleepiness).[5] Sleep/circadian disruption increases with age and cognitive impairment and even predicts preclinical AD pathology and onset of AD/ADRD. POD is associated with potential “misalignment” of human body clocks and melatonin metabolites – the key circadian hormone in sleep homeostasis. [6] Despite this, there is minimal consideration of sleep/circadian disruption in preoperative medicine. Undergoing major surgery with pre-existing sleep/circadian disruption makes it likely that these symptoms will be exacerbated during the recovery period.[7]

Our recent work and those by others have shown that suboptimal sleep/circadian regulation predicts POD risk [8–12] and progression to dementia, independent of age, sex, education, and cognition.[13] These studies also uncovered that sleep/circadian measures correlated with *CSF* amyloid/tau burden decades before dementia,[14,15] and that plasma *tau* burden was associated with PND in two surgical cohorts.[16,17] Finally, the *APOE*-ε4 genotype interacts with sleep to affect AD risk,[18] but its role in POD remains controversial.[19,20] While epidemiological evidence points to sleep/circadian regulation as a shared modifiable risk factor for POD and dementia, direct clinical evidence is lacking. Our central hypothesis is that sleep/circadian disruption promotes cognitive vulnerability after anesthesia/surgery via increased AD pathology, and this relationship is exacerbated by genetic risk for AD (see **Fig. 1** for the conceptual model).

**Fig 1.**
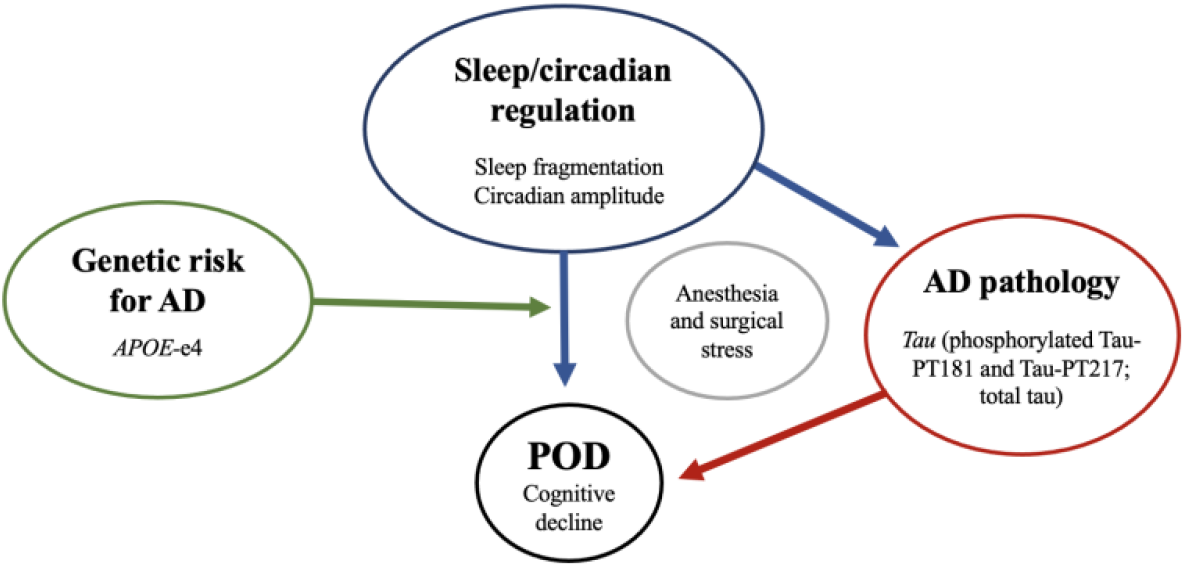
SLEEP-POD Conceptual Model: Sleep/circadian disruption effects on PND via plasma AD pathology burden and influence of genetic risk of AD. *<u>POD:</u>* postoperative delirium, *<u>APOE-ε4</u>*: apolipoprotein *ε4*.

To test this, we propose the SLEEP-POD study, a single-site, prospective, noninvasive observational study of 150 older patients (≥70y) undergoing elective, major orthopedic surgery at the Massachusetts General Hospital (MGH). This study will assess preoperative sleep and circadian rest/activity rhythms for 7 days using objective actigraphy measures and validated sleep questionnaires. Cognition and genotyping data will be collected, and *tau* (phosphorylated Tau-PT181 and Tau-PT217; total *tau*) will be measured using our published nanotechnology platform.[16,17] The primary outcome will be the incidence of POD before discharge or within 3 days of surgery.

## METHODS AND ANALYSIS

### Study design

This protocol presents a single-site, prospective, noninvasive observational study designed to examine the relationship between sleep/circadian regulation and POD and how this association is influenced by *tau* and genetic risk for AD. Subjects who meet the inclusion criteria and no exclusion criteria will be recruited after obtaining written informed consent. Recruitment will take place at the Massachusetts General Hospital (MGH). A trained study staff member will carry out the process of consenting patients. The study protocol and assessments to be performed are presented in **Table 2**.

### Study registration

This study is approved by the Institutional Review Board (IRB) at MGH, Boston. This study is registered with the US National Institutes of Health on ClinicalTrials.gov as of 19^th^ September 2023. The study will be recruiting in late September 2023 and is expected to be open for recruitment for the next 3 years (August 2026).

### Patient and public involvement

At this time, patient advisers were not involved in this research study’s design, conduct, reporting, or dissemination plans. The study is registered on ClinicalTrials.gov and updated at regular intervals. Results will be reported in a time-compliant manner on ClinicalTrials.gov.

### Inclusion and exclusion criteria

We will include 150 older (>70 years) elective orthopedic surgical patients undergoing joint replacement or spine surgery at MGH with an expected postoperative recovery of greater than 24 hours. We will include those with sufficient vision, hearing, and English language proficiency to consent and complete testing. Exclusion criteria: known dementia or related treatment, alcohol/drug abuse within 2 years, inability to wear an actiwatch, need for urgent/emergent surgery or surgery in the prior month, and more than 2-day ICU stay in the prior month. A full listing of trial exclusions and rationale is found in **Table 1**.

**Table 1.** Inclusion and exclusion criteria with rationale.

| **Table 1.** Inclusion and exclusion criteria with rationale |  |
| --- | --- |
| **Inclusion Criteria** | **Rationale** |
| ≥70 years of age | POD most commonly appears in older patients |
| Scheduled to undergo major orthopedic surgery (total hip replacement, total knee replacement, spine surgery) | Major orthopedic surgeries are associated with an increased risk for POD |
| **Exclusion Criteria** | **Rationale** |
| Inability to give consent (due to, but not limited to, preexisting dementia/MCI, cognitive impairment, severe neurocognitive damage, history of psychiatric illness, blindness, or deafness) | Inability to ethically enroll individuals of this population |
| Expected postoperative recovery for less than 24 hours | We will be unable to assess for POD |
| Patient with more than two days spent in the ICU during the month prior to surgery | Pre-existing operations/illnesses may confound the study |
| Patient with inability to wear actiwatch (due to, but not limited to, infection at the site of watch, allergies to the material of watch, limited mobility) | Required for activity-rhythm assessment |
| Non-English speaking | Inability to administer cognitive assessments that are primarily validated in English |

#### Objective drop criteria

Scheduled for a second surgical procedure during hospital stay, postoperative intubation >12 hours, or positive SARS-CoV-2 test after subject enrollment.

### Study outcome measures

#### Primary outcome

The primary outcome for this study is the incidence of delirium up to day 3 after major orthopedic surgery. Delirium will be assessed using the Confusion Assessment Method (CAM), which will be scored by the study team members twice daily on postoperative days 1-3. The CAM considers performance on attention, orientation, and memory tests from the Montreal Cognitive Assessment (MoCA) and responses to the Delirium Severity Index (DSI) – a symptom severity questionnaire. Trained study staff will use these data to complete the long CAM, which will inform a diagnostic algorithm that determines delirium status at time of assessment. If patients are discharged from the hospital before postoperative day 3, CAM evaluations up until discharge will be sufficient for delirium incidence screening.

#### Secondary outcomes

The secondary outcomes of this study include the following:

1. Severity of delirium will be measured using the CAM up until postoperative day 3 or discharge, whichever is later.
2. CAM will also measure delirium-free days during this postoperative in-hospital period.
3. Postoperative cognitive status will be recorded via telephone 1 month, 3 months, and 12 months after surgery using the Telephone Montreal Cognitive Assessment (tMoCA).
4. Postoperative health-related quality of life will be measured using a series of PROMIS questionnaires that evaluate global health (SF V.1.1), physical function (SF 8b V.1.2), pain interference (SF 8a V.1.0), applied cognition abilities (SF 8a V.1.0), and sleep disturbance (SF 4A V.1.0) at 1 month, 3 months, and 12 months after surgery.
5. Associations with baseline and changes in AD-related pathology (phosphorylated Tau-PT181 and Tau-PT217; total *tau*) will be inferred from blood samples taken prior to surgery and once on POD 1. Nutrition assessments will be performed prior to all blood collections.
6. To gather information about AD variant genes, including APOE-ε4, we will send de-identified blood samples for genotyping.

#### Other outcomes

Rest-activity rhythm data from accelerometer watches will be assessed as an exploratory outcome measure. The watch will collect data including sleep duration, regularity, stability, and timing of activity rhythms. Patients will return the watch on the day of surgery and will be asked to wear it for an additional week starting the day of their one-month, 3-month, and 12-month follow-up visits. Patients will be able to access the results that we derive from the actiwatch data. These results will include information about their sleep habits and circadian rhythm.

In addition, patients will be evaluated during the baseline interview with continuous ECG recording to explore vascular biomarkers associated with delirium. A 4-lead ECG with oximetry will be placed on the patients by trained study staff during the inclusion visit. If any abnormalities appear in the patient’s ECG that they were unaware of, they will be advised to contact their doctor. We will explore pulse wave velocity (PWV) and augmentation index (AI) to evaluate artery stiffness and peripheral resistance, given potential relationships to sleep health and delirium.

#### Clinical Data Collection

Clinical data including age, sex, body mass index (BMI), comorbidities, length of hospital stay, and surgical details (e.g., surgery duration, anesthesia type, blood loss, complications) will be recorded using passively collected data from the patient’s electronic medical record.

### Data collection

**Table 2.** Study event schedule.

| **Table 2. Study event schedule** |  |  |  |  |  |
| --- | --- | --- | --- | --- | --- |
|  | **Screen**<br>(<3 months) | **Preoperative**<br>(<6 weeks) | **Postoperative**<br>(1-3 days) | **Follow-Up**<br>(1 month) | **Follow-Up**<br>(3 & 12 months) |
| Informed Consent | X |  |  |  |  |
| eMR Data Collection* | X | X | X | X | X |
| Inclusion/ Exclusion | X |  |  |  |  |
| CAM Evaluation* |  |  | X | X | X |
| MoCA/tMoCA* |  | X | X | X | X |
| PROMIS<br>Questionnaires** |  | X | X | X | X |
| Blood Collection |  | X | X |  |  |
| Nutrition Assessment |  | X | X |  |  |
| Actiwatch Data<br>Collection (1-week) |  | X |  | X |  |
| ECG (PWV, AI)* |  | X |  |  |  |

\* *eMR* = electronic medical record, *CAM* = Confusion Assessment Method, *MoCA/tMoCA* = (telephone) Montreal Cognitive Assessment, *ECG (PWV, AI)* = electrocardiogram (pulse wave velocity, augmentation index)
\*\* *PROMIS* = Patient-Reported Outcomes Measurement Information System: physical function (SF 8b V.1.2) and applied cognition abilities (SF 8a V.1.0) may only be evaluated during follow-up visits.

### Recruitment

Clinical research coordinators will identify potential subjects presenting to Massachusetts General Hospital for orthopedic surgery. Whenever possible, a member of the study team (clinical research coordinator, principal investigator, or co-investigator) will meet the patient during the orthopedic surgery clinic visit prior to surgery. The study procedures will be explained in detail at this time, and consent may occur. Eligible patients whom we cannot connect with at their preoperative clinic visit will be contacted via telephone.

Previous studies have shown that POD affects older patients (>70 years). In addition, on the rare occasions when patients develop POD at younger ages (<70), they are more likely to represent a scientifically distinct population. Thus, we will focus our research question on patients over 70 to obtain unbiased and scientifically valid results. We will enroll 150 patients for this study. We anticipate that our study will include women and minority populations representative of the Mass General Brigham and our regional location in Suffolk County, Boston, MA.

### Data analysis

We will use logistic regression for a dichotomized POD outcome to examine sleep/circadian predictors. We will then use multivariate linear regression to test our continuous exposures (sleep fragmentation and circadian amplitude) and continuous outcomes (e.g., cognition). For mediation testing, we will bootstrap the indirect effect CIs, the optimal method from Fritz & MacKinnon’s work,^6^ which would provide >80% power to detect the medium-medium-zero effect sizes condition (α = 0.39, β = 0.39, τ′ = 0). There cannot be mediation if no associations between sleep and *tau* exist. We will then test whether *tau* burden interacts with sleep/circadian regulation associated with POD. We will introduce interaction terms between APOE-ε4 status and our two exposure variables into our first model.

The model fit, and outlier assessment will be examined using formal fit criteria and model inspection. We will select covariates based on prior POD or AD studies and the strength of association/extent of confounding while avoiding model overfitting. We will assess a core model with age and sex and build an adjusted model with the optimal number of predictors after evaluating with the Akaike Information Criterion (AIC).

### Data management and quality assurance

This study will employ a web-based portal for data quality and completeness. The portal will display in real-time the following variables for all patients: sex, race, adverse events, study-related data, etc. To ensure data is accurately and completely collected during the study, the PI will be responsible for ensuring the study protocol is being followed, the IRB has approved changes to the protocol, and all facilities are appropriate for the conduct of the study. Also, the PI will review subject records to determine whether the data collected is accurate, complete, and current. A Scientific Review will occur annually and consist of a review of subject recruitment, staff training, and quality control procedures. Monitoring progress reports will be submitted to regulatory and/or funding bodies (IRB, Alzheimer’s Association) as requested and/or required.

### Limitations

Limitations of the study include its observational nature and, therefore, the inability to demonstrate causation. Treatment effects after elective joint surgery may affect the association between sleep/circadian rhythms and delirium. In addition, repeat follow-up 12 months after surgery may increase the likelihood of missing data.

Despite these pitfalls, older orthopedic patients have similar characteristics, which mitigates variability within this study. They also have a high co-morbid burden of sleep disturbances, which can enrich the signal for POD and cognitive decline.

Patient burden during the study could originate from discomfort while wearing the actiwatch, irritation at IV insertion, irritation when ECG pads are removed, and fatigue during cognitive assessments. When possible, blood collection will be performed while the patient is under general anesthesia and extracted from an indwelling line to minimize patient discomfort. All other efforts will be made to ensure minimal patient discomfort and stress throughout the study.

### Ethics and dissemination

The principal investigator of this study will be responsible for final decisions regarding changes to the protocol. The PI will report such changes to the MGB IRB. Electronic patient information will be accessed only as necessary throughout the completion of the study. All of the data collected from the study will be accessible to the PI. The main papers reporting information about this study will present primary and secondary outcomes. Additional analyses will be performed to formulate predictive models for delirium using sleep/circadian patterns and genetic data. Manuscripts describing the mechanisms for delirium pathophysiology from substudies (i.e., circadian/sleep disturbance, actigraphy, protein biomarkers) will also be published. Plans for dissemination include conference presentations at a variety of scientific institutions. Results from this study are intended to be published in peer-reviewed journals. Relevant updates will be made available on ClinicalTrials.gov.

In conclusion, this study will investigate mechanisms for delirium through analysis of sleep/circadian disruption and AD pathology. We present a novel approach to understanding a modifiable risk factor for postoperative delirium and cognitive decline after surgery.

## Data Availability

All data produced in the present study will be available upon reasonable request to the authors.

## Acknowledgments

The authors would like to acknowledge all members involved in data collection and administration.

## Author Statement

E.S., P.L., A.M., E.M., O.A., Z.X., K.H., and L.G. were responsible for conceptualising trial design. E.S., E.G., A.M., and L.G. are responsible for recruitment, enrollment, and data collection. E.S., E.G., S.N.B., P.L., C.G., A.M., H.D., S.S., E.F.G., R.S., E.M., O.A., Z.X., K.H., and L.G. have critically revised the protocol and approved the final version.

